# Variation at *COMT, ADH1B-ADH1C* and *HTR2A* is associated with substance use disorders in Ukrainians

**DOI:** 10.64898/2026.04.23.26351594

**Authors:** Vitalina Bashynska, Oksana Zahorodnia, Yuliia Borysovych, Yaroslav Zaplatnikov, Valeriia Vasylieva, Ihor Arefiev, Nariman Darvishov, Dariia Osychanska, Arthur Karapetov, Olena Melnychuk, Olena Boiko, Gennadii Zil’berblat, Olena Turos, Inga Prokopenko, Marika Kaakinen

**Affiliations:** Laboratory of Biocides and Toxicology, State Institution «Marzieiev Institute for Public Health of the National Academy of Medical Sciences of Ukraine», 50 Hetman Pavlo Polubotka Street, 02094, Kyiv, Ukraine; Ukrainian Institute for Systems Biology and Medicine, Dorogozhitska Str. 3, 04119, Kyiv, Ukraine; Educational Scientific Institute of High Technologies, Taras Shevchenko National University of Kyiv, Kyiv, Ukraine; Educational And Scientific Center “Institute of Biology and Medicine”, Taras Shevchenko National University of Kyiv, Kyiv, Ukraine; Faculty of Psychology, Taras Shevchenko National University of Kyiv, Kyiv, Ukraine; MedLux Rehabilitation Center, Kyiv, Ukraine; Regional Psychiatric and Narcologic Medical Association, Glevakha, Ukraine; Laboratory of Air Quality, State Institution «Marzieiev Institute for Public Health of the National Academy of Medical Sciences of Ukraine», Kyiv, Ukraine; Department of Clinical and Experimental Medicine, School of Biosciences, University of Surrey, Guildford, UK; People-Centred Artificial Intelligence Institute, University of Surrey, Guildford, UK; Department of Epidemiology and Biostatistics, School of Public Health, Imperial College London, London, United Kingdom; Institute for Molecular Medicine Finland, University of Helsinki, Helsinki, Finland

**Keywords:** substance use disorders, genetic association study, Ukraine, *ADH1B-ADH1C*, *HTR2A*, *COMT*

## Abstract

**Background:** Substance use disorders (SUDs), including alcohol and drug dependence, and smoking, pose a public health threat with their high prevalence and comorbidity with other diseases, and contribution to mortality. SUDs are highly correlated, and their genetic background is shared to some degree.

**Objectives:** We aimed to investigate the genetic associations of previously reported loci for a wide range of SUDs in an unstudied Ukrainian population.

**Methods:** We collected data from 595 individuals (339 women, 253 men), including 321 participants from two rehab centres. Based on clinical review and questionnaire data we defined drug dependence, alcohol dependence, alcohol abuse, binge drinking, smoking, opiate, amphetamine, cannabis, and hallucinogen use, along with several intermediary alcohol use and smoking variables considering the amount of use and the level of dependence. We genotyped *COMT*-rs4680, *ADH1B-ADH1C*-rs1789891, and *HTR2A*-rs6313, and applied logistic and ordered logistic regression assuming an additive inheritance model, controlling for the recruitment group, other substance uses, age, and sex, in the association analyses.

**Results:** We replicate (*P*<0.05) the associations at *COMT*-rs4680 with smoking status (OR[95% CI]=1.56[1.01-2.41], *P*=0.047) and heaviness (1.37[1.04-1.80], *P*=0.026), and at *ADH1B-ADH1C*-rs1789891 and *HTR2A*-rs6313 with alcohol dependence (1.69[1.03-2.76], *P*=0.038 and 0.66[0.47-0.92, *P*=0.016], respectively). Furthermore, we provide evidence for an association at *HTR2A*-rs6313 with hallucinogen use (0.58[0.35-0.98], *P*=0.040).

**Conclusion:** In this study on multiple SUDs we shed light on the genetic background of SUDs in Ukrainians and provide further evidence that variation at *COMT* is mainly associated with smoking, at *ADH1B-ADH1C* with alcohol-related variables, whereas *HTR2A* is a more general SUD-associated locus.

**Highlights:**

- We present the first genetic study of substance use disorders in Ukrainians
- We replicate the associations at *COMT*-rs4680 with smoking status and heaviness, and *ADH1B-ADH1C*-rs1789891 and *HTR2A*-rs6313 with alcohol dependence
- We provide evidence for an association at *HTR2A*-rs6313 with hallucinogen use

## 1. Introduction

Substance use disorders (SUDs), including drug dependence (DD), alcohol dependence (AD), and smoking, are a global threat to public physical and mental health. Their estimated prevalence lies within a broad range from ∼4% for drug use disorder to >20% for smoking (WHO, 2017; Ritchie, Roser, 2018; UNODC, 2023). They are highly comorbid with and substantially increase risk for multiple diseases, including other psychiatric and metabolic disorders, as well as cancers (Grant et al, 2016; Zima 2018; Park et al. 2023). Each of them annually leads to millions of deaths worldwide (Deak, Johnson, 2021). SUDs particularly affect low- and middle-income countries (El Hayek et al. 2024). Particularly, in Ukraine, the prevalence of alcohol use disorder in 2021 was 3.6%, and the country ranked 6th among countries for alcohol-attributable fractions of all-cause deaths (Ritchie, Roser, 2018). According to the Global Adult Tobacco Survey (GATS), in 2017, 20.1% of the adult population in Ukraine smoked daily (WHO, 2017). UN data in 2019 reported the prevalence of amphetamine use in Ukraine at the level of 0.3%, and for opiates it was 1.05% (UNODC, 2019).

Twin studies suggest that genetic variation accounts for 30-70% of SUDs variability, depending on substance (Ducci, Goldman, 2012). Nevertheless, the understanding of the genetics of SUDs is still far behind that of other complex diseases. Genome-wide association studies (GWASs) of SUDs carried out to date have reported at a maximum of tens of loci at genome-wide significance (Zhou et al., 2020; Gerring, et al. 2024), in contrast to other disorders, where hundreds of independent risk loci are established (Cotsapas, Mitrovic, 2018; Cerezo, et al. 2025). Polygenic scores (PGS) created for some SUD phenotypes, e.g. AD, explain only a small proportion of phenotypic variation (Savage et al. 2018), and their clinical utility can still be improved (Kember et al, 2024).

Emerging evidence suggests that psychiatric traits are better understood as a spectrum than as a set of distinct nosologies (Adam 2013). SUDs, being part of the broader domain of behavioural and mental disorders, represent a set of epidemiologically correlated phenotypes – both among themselves (Kember et al. 2024) and with other psychiatric conditions, including schizophrenia, major depressive disorder, and post-traumatic stress disorder (Wu et al, 2018, Gerring, et al. 2024). Nevertheless, different SUDs may be associated with partially distinct genetic risk factors, as demonstrated for alcohol dependence, alcohol consumption levels, binge drinking and problematic alcohol use (Zhou et al. 2020; Deak, Johnson, 2021; Sanchez-Roige et al. 2021). This motivates to study a wide set of SUDs within the same population to disentangle their shared and distinct genetic risk factors.

Genetic associations with complex polygenic traits can differ in populations with different evolutionary histories (Marigorta et al. 2011; Araújo, Wheeler, 2022). For instance, PGSs for different SUDs and other psychiatric traits show limited transferability between different ancestries (Gerring, et al. 2024; Hartwell et al. 2024). In line with this, a recent population-based genetic study has revealed significant differences between a pilot dataset of Ukrainians and Russian and Central European populations (Oleksyk et al. 2021): the whole-genome sequencing data for these populations do not cluster together on the principal component analysis plots. Moreover, sequencing of Ukrainian genomes has allowed to identify 478,000 novel genomic SNPs that have never been registered in the Genome Aggregation Database (gnomAD) (Karczewski et al. 2020). This highlights the need to improve our knowledge on the genetic architecture of different phenotypes, including SUDs, in Ukrainians.

Here, we present the results of the first genetic association study with multiple SUDs, including AD, DD, smoking and related phenotypes in Ukrainians. We aimed to investigate the nature of associations with SUDs in Ukrainians at four loci which have previously been implicated in GWASs or consistently replicated in candidate gene studies for SUDs in other populations: *COMT*-rs4680, *ADH1B-ADH1C*-rs1789891, *HTR2A*-rs6313, and *OPRM1*-rs1799971. Each of these loci has been associated with at least two SUDs in GWASs or candidate gene studies (**Supplementary Table A1**). For instance, *ADH1B-ADH1C*-rs1789891 was the top GWAS-identified signal for AD that had been replicated in follow up studies at the time of inclusion (Frank et al. 2012; Treutlein et al. 2017).

## 2. Material and methods

### 2.1. Study population

The study included 595 volunteers (339 women, 253 men) living in Ukraine, with a mean age of 32.5±10.7 years (range 16 – 76 years). Of them, 321 participants were undergoing rehabilitation in one of the available rehab centres of Kyiv region: MedLux rehab centre or Kyiv Regional Psychiatric and Narcological Medical Association in Glevakha. The remaining 274 participants were recruited via e-mails, social media posts, or personal invitations to take part in a substance use research. All the participants donated blood or buccal epithelium samples for DNA extraction, and responded to a standardised questionnaire, which included the Barratt impulsivity scale (Patton et al. 1995), cigarette dependence scale (CDS-5) (Etter et al. 2003), a part of the Alcohol Use Disorders Identification Test (AUDIT-C) that characterises regular alcohol consumption (Bush et al. 1998; Bradley et al. 2003), a supplementary clinical questionnaire focusing on “Cutting down”, “Annoyance by criticism”, “Guilty feeling”, and “Eye-openers” (CAGE) (Ewing et al. 1984; Brown, Rounds, 1995) as well as several other questions related to their lifestyle, substance use preferences, social, and demographic background.

Interpretation of the standardised scales included in the questionnaire was carried out in accordance with their standard guidelines. In addition, for the rehab patients, their medical history records relevant to their substance use, were provided and reviewed by professionals at the rehab centres.

All the participants gave written informed consent. The study was approved by the local ethics committees (Record No 8 From 14 August 2020, Institute of Gerontology, NAMS Ukraine; Record N 1 from 11.06.2024, O.M. Marzieiev Institute for Public Health NAMS Ukraine), and by the University of Surrey Ethics Committee (EGA ref: FHMS 20-21 198 EGA, approved on 7 October 2021).

### 2.2. Phenotype definition

### 2.2.1. Binary phenotypes

The individuals’ case/control statuses for each phenotype were defined depending on their ICD-10 diagnosis, when available, and the output from the questionnaire, and are described in **Table 1**. The phenotype ‘drug dependence (DD)’ was assigned exclusively to people who had an ICD-10 diagnosis, set by a qualified narcologist, of “F11/F15/F19” (Mental and behavioural disorders due to use of opioids/stimulants/multiple drug use and use of other psychoactive substances, correspondingly), whereas ‘alcohol dependence (AD)’ required an ICD-10 diagnosis code of “F10.2” (Mental and behavioural disorders due to use of alcohol. Alcohol dependence). The people outside rehab centres might not have a diagnosis for AD or for the broader alcohol use disorder (AUD) but based on the AUDIT-C and CAGE questionnaires could be considered heavy drinkers. Therefore, we defined a variable describing a problematic drinker: ‘alcohol abuse (AA)’, a complex phenotype which represents either an ICD-10 diagnosis “F10.2” or being an excessively high drinker with signs of self-control problems, i.e. drinking at least 5-6 drinks at least 2-3 times per week based on the questionnaires AUDIT-C and CAGE.

**Table 1.** Sample characteristics.

| Phenotype | Based on | Levels | Case definition | Control definition | Cases, N (%) | Controls, N(%) | N(%) missing |
| --- | --- | --- | --- | --- | --- | --- | --- |
| **Drugs** |  |  |  |  |  |  |  |
| Drug dependence (DD) | ICD-10 F11/F15/F19 | yes/no | ICD-10 F11/F15/F19 diagnosis | No ICD-10 F11/F15/F19 diagnosis | 172 (28.9) | 390 (65.5) | 33 (5.5) |
| Opiate dependence | ICD-10 F11 | yes/no | ICD-10 F11 diagnosis | No ICD-10 F11 diagnosis | 92(15.5) | 459 (77.1) | 44 (7.4) |
| Amphetamine dependence | ICD-10 F15 | yes/no | ICD-10 F15 diagnosis | No ICD-10 F11 diagnosis | 145 (24.4) | 415 (69.7) | 35 (5.9) |
| Cannabis use | Questionnaire | yes/no | Self-reported use | Self-reported never used | 174 (29.2) | 232 (39.0) | 189 (31.8) |
| Hallucinogen use | Questionnaire | yes/no | Self-reported use | Self-reported never used | 66 (11.1) | 306 (51.4) | 223 (37.5) |
| **Alcohol** |  |  |  |  |  |  |  |
| Alcohol abuse (AA_ctrl\|STRICT) | ICD-10 F10.2 or AUDIT-C and CAGE questionnaire | yes/no | ICD-10 F10.2 or at least 5-6 drinks at least 2-3 times per week | 0-2 on AUDIT-C regular consumption component and 0-1 on CAGE | 203 (34.1) | 186 (31.3) | 206 (34.6) |
| Alcohol abuse (AA_ctrl\|RELAXED) | ICD-10 F10.2 or AUDIT-C and CAGE questionnaire | yes/no | ICD-10 F10.2 or at least 5-6 drinks at least 2-3 times per week | 0-3 on AUDIT-C regular consumption component and 0-2 on CAGE | 203 (34.1) | 247 (41.5) | 145 (24.4) |
| Alcohol abuse (AA3level) | ICD-10 F10.2 or AUDIT-C and CAGE questionnaire | yes/intermediate level/no level | Cases: ICD-10 F10.2 or at least 5-6 drinks at least 2-3 times per week; Intermediate level: in-between these patterns, excluding missing or inconsistent data | 0-3 on AUDIT-C regular consumption component and 0-2 on CAGE | 203 (34.1)/91 (15.3) | 247 (41.5) | 54 (9.1) |
| Alcohol dependence (AD_ctrl\|STRICT) | ICD-10 F10.2 | yes/no | ICD-10 F10.2 | 0-2 on AUDIT-C regular consumption component and 0-1 on CAGE | 152 (25.5) | 186 (31.3) | 257 (43.2) |
| Alcohol dependence (AD_ctrl\|RELAXED) | ICD-10 F10.2 | yes/no | ICD-10 F10.2 | 0-3 on AUDIT-C regular consumption component and 0-2 on CAGE | 152 (25.5) | 247 (41.5) | 196 (32.9) |
| Alcohol dependence (AD3level) | ICD-10 F10.2 | yes/intermediate level/no level | Cases: ICD-10 F10.2; Intermediate level: in-between these patterns, excluding missing or inconsistent data | 0-3 on AUDIT-C regular consumption component and 0-2 on CAGE | 152 (25.5)/142 (23.9) | 247 (41.5) | 54 (9.1) |
| AUDIT9level | AUDIT-C | 9 levels | A sum score from the two AUDIT-C components on how often ('never/ 'no more often than once per month', '2-4 times per month', '2-3 times per week', 'more often than 3 times per week') and what amount of alcohol (0-2, 3-4, 5-6, 7-9, >10 units of alcohol at the time of consumption) the participants used | NA | 25(4.2)/19(3.2)/35(5.9)/53(8.9)/72(12.1)/86(14.4)/117(19.7)/103(17.3)/ 32(5.4) |  | 53(8.9) |
| Binge drinking | AUDIT-C and medical history reports | yes/no | Drinking 6 drinks and more at least once a month or severe drinking episodes without being sober for days in self-report or medical history | The declared absence of this drinking pattern | 148<br>(24.9) | 317<br>(53.3) | 130<br>(21.8) |
| **Smoking** |  |  |  |  |  |  |  |
| Smoking, ever | CDS-5 | yes/no | >5 packs of cigarettes/lifetime | <=5 packs of cigarettes/lifetime and never a regular smoker | 397<br>(66.7) | 162<br>(27.2) | 36(6.1) |
| Smoking,<br>current/former<br>(Smoking3level) | CDS-5 | current/<br>former/<br>never | Current: >5 packs of<br>cigarettes/lifetime<br>and smoking<br>currently; Former: >5<br>packs of<br>cigarettes/lifetime<br>and not smoking<br>currently | <=5 packs of<br>cigarettes/lifeti<br>me and never<br>a regular<br>smoker | 307<br>(51.6)/90<br>(15.1) | 162<br>(27.2) | 36 (6.1) |
| Cigarettes per<br>day (CPD5level) | CDS-5 | 5 levels | 0-5, 6-10, 11-20, 21-<br>29, 30+ | NA | 18(3.0)/39(6.6)/<br>167(28.0)/66(11.1)/<br>221(37.1) |  | 84(14.1) |
| Smoking<br>dependence<br>(CDS3level) | CDS-5 | 3 levels | Breaks at values 5,<br><15, <26 in<br>accordance with the<br>distribution of the<br>data | NA | 183<br>(30.8)/<br>175<br>(29.4) | 210<br>(35.3) | 27(4.5) |

AA\_ctrlSTRICT, alcohol abuse/problematic drinking, 'strict' control definition; AA\_ctrlRELAXED, alcohol abuse/problematic drinking, 'relaxed' control definition; AA, case/intermediate phenotype – 3-level phenotype on alcohol abuse/problematic drinking; AD\_ctrlSTRICT, alcohol dependence, 'strict' control definition; AD\_ctrlRELAXED, alcohol dependence, 'relaxed' control definition; AD, case/intermediate phenotype, 3-level phenotype on alcohol dependence.

For assignment of the control status in alcohol-related phenotypes, two cutoffs of the scales were used. ‘Strict’ controls scored 0-2 on AUDIT-C regular consumption component and had low scoring (0-1) on CAGE. ‘Relaxed’ controls had AUDIT-C regular consumption scoring 0-3 (median) and 0-2 scoring on CAGE (Brown, Rounds, 1995; Verhoog et al. 2020). During assignment of status, all the questionnaires had been checked for inconsistencies in answers. In addition to this, we checked that the controls did not report problems with alcohol in their medical histories.

‘Binge drinking’ phenotype was defined separately based on the results from the AUDIT-C and medical history reports, in accordance with standard criteria (NIAAA, 2025).

Phenotype ‘smoker’ was assigned to people who self-reported having smoked more than five packs of cigarettes in a lifetime (Marees et al., 2018) and being a current or former smoker. People who had smoked less and did not report themselves as ever regular smokers in the CDS-5 questionnaire, were considered controls.

Case and control phenotypes for cannabis and hallucinogen use were assigned based on their questionnaire responses.

#### 2.2.2. Multi-level categorical phenotypes

Given the complexity of drinking and smoking phenotypes and many participants with intermediate phenotypes in relation to the above-mentioned substances, we introduced phenotypes that would characterise an intermediate level of dependence or better describe the broad distribution of smoking and alcohol use patterns.

For smoking, we have split the smoker phenotype to current, former and never smokers (Smoking3level). We further categorised cigarettes per day (CPD) obtained from the CDS-5 questionnaire into five levels (CPD5level): 0-5, 6-10, 11-20, 21-29, and 30+ CPDs. In addition, the whole CDS-5 scale measuring smoking dependence was categorized (CDS3level) into equal-sized groups with breaks at 5, 15 and 26 (**Supplementary Figure A1, Table 1**).

For alcohol use, in addition to the above-mentioned case/control levels, we have introduced an intermediate level of AD and AA phenotypes (AD3level and AA3level, respectively), which included people that have an AUDIT-C regular consumption component scoring above recommended cut-offs for controls (>3) (Reinert, Allen, 2007; Frank et al. 2008), and/or tend to lose control of the drinking amount, while not having an obvious problematic use, i.e. having 6 and more portions of alcohol once a month. We further obtained a categorical variable ‘AUDIT9level’ as a sum score from the nine levels of the AUDIT-C questionnaire on how often (‘never’/’no more often than once per month’, ‘2-4 times per month’, ‘2-3 times per week’, ‘more often than 3 times per week’) and what amount of alcohol (0-2, 3-4, 5-6, 7-9, >10 units of alcohol at the time of consumption) the participants used. The distributions of the multi-level categorical phenotypes are shown in the **Supplementary Figure A1**.

### 2.3. DNA extraction and genotyping

DNA was extracted using Quick-DNA Miniprep (ZYMO RESEARCH). Genotyping of the selected four loci, *COMT* rs4680, *ADH1B-ADH1C* rs1789891, *HTR2A* rs6313, and *OPRM1* rs1799971, was carried out using custom-made primers and TaqMan probes (**Supplementary Table A1**). After the completion of genotyping, ∼20% of samples were genotyped in duplicates to ensure genotyping accuracy. Hardy-Weinberg equilibrium of the genotypes was checked using χ2 test in people outside rehab centres (n=272) and additionally in those who were further non-smokers (n=143). The genotype at rs1799971 *OPRM1* did not pass the Hardy-Weinberg equilibrium test in either of the tested sets and was therefore excluded from the genetic association analysis.

### 2.4. Statistical analysis

The analyses were performed using the R and RStudio software (R Core Team, 2025; Posit team, 2025).

We have calculated the Spearman correlation between the phenotypes and compared the genotype distributions among Ukrainian people outside the rehab centres with the EUR population in the 1000 Genomes project (2015) using χ2 test. For the genetic association analysis with binary outcomes (DD, AD, AA, and specific drug use), we used logistic regression, and with multi-level categorical outcomes (AUDIT9level, CDS8level, CPD5level, Smoking3level, AA3level, AD3level) we used ordered logistic regression. We used an additive genetic model testing for associations with the outcome and the increase in each copy of the effect allele. Given the considerable co-occurrence of substance use phenotypes in our sample (**Figure 1A**), all analyses were adjusted for the remaining substance phenotypes, as well as age, sex, and the recruitment group, i.e. people outside rehab centres, people from MedLux rehab and people from Glevakha rehab. The substance phenotype covariates varied for different analyses and are described in **Table 2**. The results are presented as odds ratios (OR) with their 95% confidence intervals (CI) and *P*-values. A statistically significant association was considered at a Bonferroni-corrected threshold of *P*<0.017 to account for three tested SNPs, while a suggestive association was considered at a nominal significance level of *P*<0.05.

**Figure 1A.**
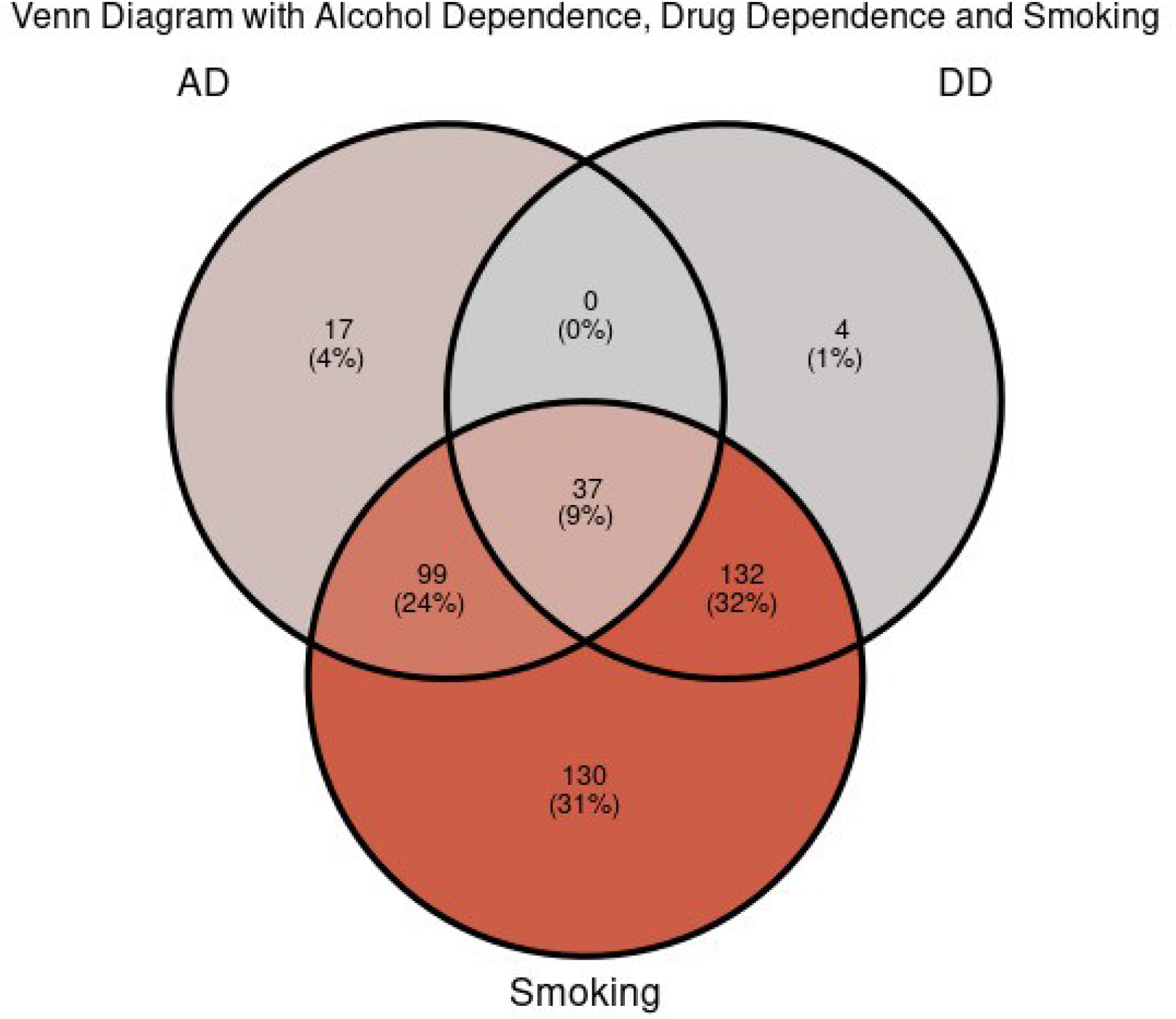
Venn diagram showing co-occurrence of the major substance use phenotypes in the sample: alcohol dependence (AD), drug dependence (DD), and smoking.

**Table 2.**
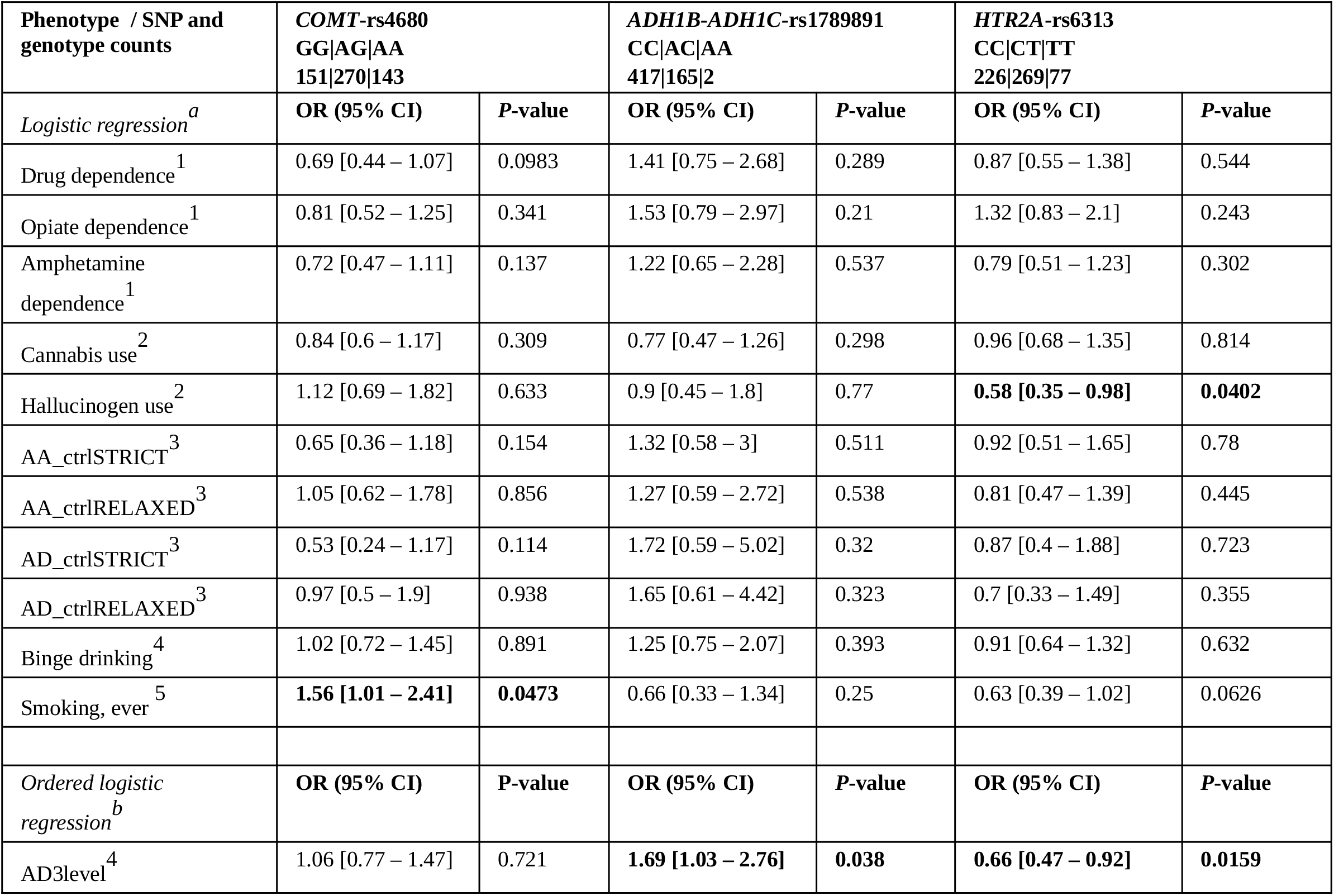
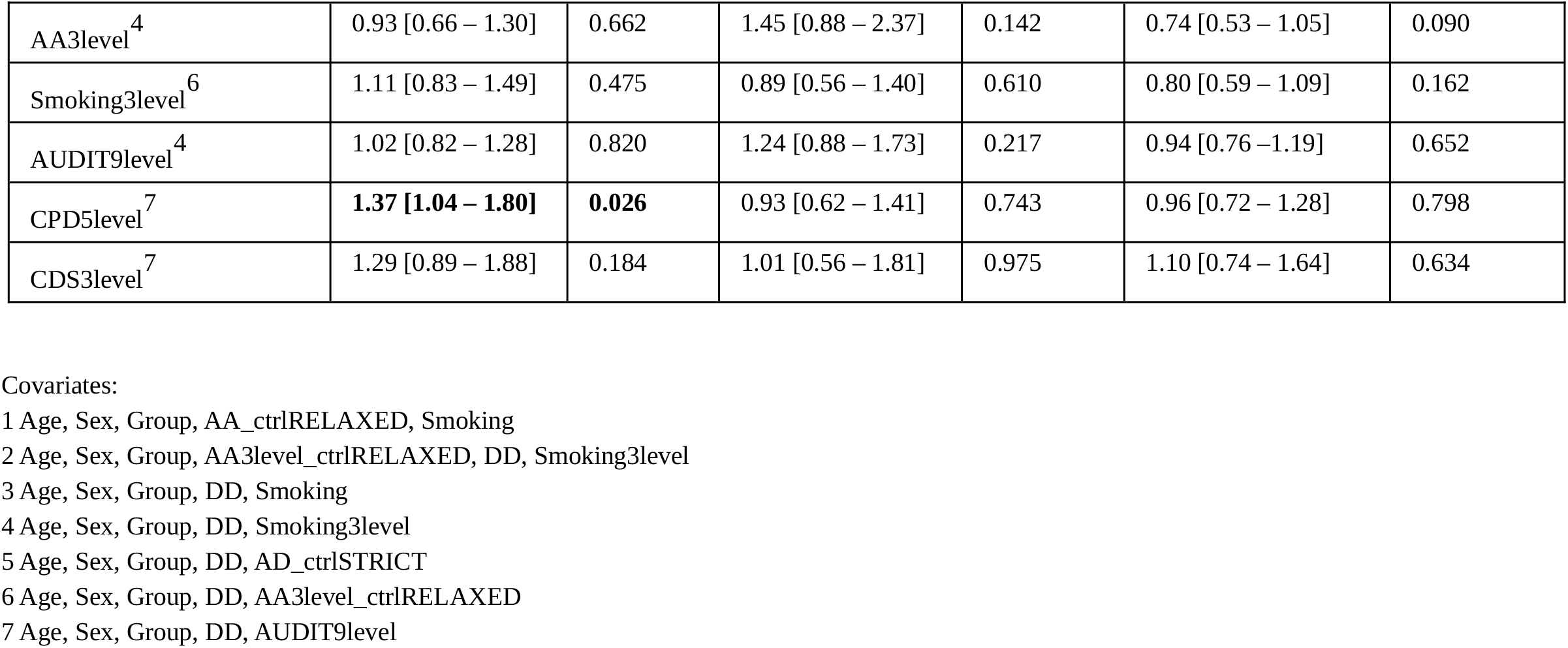
Results from the logistic and ordered logistic regression between each of the SUD phenotypes and *COMT-*rs4680, *ADH1B-ADH1C-*rs1789891, and *HTR2A*-rs6313.

| Phenotype / SNP and genotype counts | *COMT*-rs4680<br>GG\|AG\|AA<br>151\|270\|143 |  | *ADH1B-ADH1C*-rs1789891<br>CC\|AC\|AA<br>417\|165\|2 |  | *HTR2A*-rs6313<br>CC\|CT\|TT<br>226\|269\|77 |  |
| --- | --- | --- | --- | --- | --- | --- |
|  | OR (95% CI) | P-value | OR (95% CI) | P-value | OR (95% CI) | P-value |
| *Logistic regression*<sup>a</sup> |  |  |  |  |  |  |
| Drug dependence<sup>1</sup> | 0.69 [0.44 – 1.07] | 0.0983 | 1.41 [0.75 – 2.68] | 0.289 | 0.87 [0.55 – 1.38] | 0.544 |
| Opiate dependence<sup>1</sup> | 0.81 [0.52 – 1.25] | 0.341 | 1.53 [0.79 – 2.97] | 0.21 | 1.32 [0.83 – 2.1] | 0.243 |
| Amphetamine dependence<sup>1</sup> | 0.72 [0.47 – 1.11] | 0.137 | 1.22 [0.65 – 2.28] | 0.537 | 0.79 [0.51 – 1.23] | 0.302 |
| Cannabis use<sup>2</sup> | 0.84 [0.6 – 1.17] | 0.309 | 0.77 [0.47 – 1.26] | 0.298 | 0.96 [0.68 – 1.35] | 0.814 |
| Hallucinogen use<sup>2</sup> | 1.12 [0.69 – 1.82] | 0.633 | 0.9 [0.45 – 1.8] | 0.77 | **0.58 [0.35 – 0.98]** | **0.0402** |
| AA_ctrlSTRICT<sup>3</sup> | 0.65 [0.36 – 1.18] | 0.154 | 1.32 [0.58 – 3] | 0.511 | 0.92 [0.51 – 1.65] | 0.78 |
| AA_ctrlRELAXED<sup>3</sup> | 1.05 [0.62 – 1.78] | 0.856 | 1.27 [0.59 – 2.72] | 0.538 | 0.81 [0.47 – 1.39] | 0.445 |
| AD_ctrlSTRICT<sup>3</sup> | 0.53 [0.24 – 1.17] | 0.114 | 1.72 [0.59 – 5.02] | 0.32 | 0.87 [0.4 – 1.88] | 0.723 |
| AD_ctrlRELAXED<sup>3</sup> | 0.97 [0.5 – 1.9] | 0.938 | 1.65 [0.61 – 4.42] | 0.323 | 0.7 [0.33 – 1.49] | 0.355 |
| Binge drinking<sup>4</sup> | 1.02 [0.72 – 1.45] | 0.891 | 1.25 [0.75 – 2.07] | 0.393 | 0.91 [0.64 – 1.32] | 0.632 |
| Smoking, ever<sup>5</sup> | **1.56 [1.01 – 2.41]** | **0.0473** | 0.66 [0.33 – 1.34] | 0.25 | 0.63 [0.39 – 1.02] | 0.0626 |
| *Ordered logistic regression*<sup>b</sup> | OR (95% CI) | P-value | OR (95% CI) | P-value | OR (95% CI) | P-value |
| AD3level<sup>4</sup> | 1.06 [0.77 – 1.47] | 0.721 | **1.69 [1.03 – 2.76]** | **0.038** | **0.66 [0.47 – 0.92]** | **0.0159** |
| AA3level<sup>4</sup> | 0.93 [0.66 – 1.30] | 0.662 | 1.45 [0.88 – 2.37] | 0.142 | 0.74 [0.53 – 1.05] | 0.090 |
| Smoking3level<sup>6</sup> | 1.11 [0.83 – 1.49] | 0.475 | 0.89 [0.56 – 1.40] | 0.610 | 0.80 [0.59 – 1.09] | 0.162 |
| AUDIT9level<sup>4</sup> | 1.02 [0.82 – 1.28] | 0.820 | 1.24 [0.88 – 1.73] | 0.217 | 0.94 [0.76 – 1.19] | 0.652 |
| CPD5level<sup>7</sup> | **1.37 [1.04 – 1.80]** | **0.026** | 0.93 [0.62 – 1.41] | 0.743 | 0.96 [0.72 – 1.28] | 0.798 |
| CDS3level<sup>7</sup> | 1.29 [0.89 – 1.88] | 0.184 | 1.01 [0.56 – 1.81] | 0.975 | 1.10 [0.74 – 1.64] | 0.634 |

Covariates:
1 Age, Sex, Group, AA\_ctrlRELAXED, Smoking
2 Age, Sex, Group, AA3level\_ctrlRELAXED, DD, Smoking3level
3 Age, Sex, Group, DD, Smoking
4 Age, Sex, Group, DD, Smoking3level
5 Age, Sex, Group, DD, AD\_ctrlSTRICT
6 Age, Sex, Group, DD, AA3level\_ctrlRELAXED
7 Age, Sex, Group, DD, AUDIT9level

## 3. Results

The sample included more men than women (57% vs. 42.5 %, 0.5% had missing sex information, **Table 1**). In this study, 28.9% individuals had ICD-10 drug dependence (DD; F11/F15/F19 diagnosis, **Table1**). Among all drugs being used by the participants, cannabis was the most frequently used drug (29.2%), followed by amphetamine (24.4%). Hallucinogen and opiate uses were less commonly reported, however, for hallucinogen use as well as for cannabis use, we had the largest missing data (**Table 1**). About a third of the sample (34.1%) were categorised as problematic drinkers (alcohol abuse, AA), whereas 25.6% of the participants had a diagnosed alcohol use disorder (alcohol dependence, AD) and in the same proportions were the binge drinkers (24.9%) in the sample. Smoking was common with 66.7% reporting to smoke currently or previously.

Significant Spearman correlations between the studied phenotypes are presented in **Figure 1B**, with the strongest correlations observed among phenotypes within the same substance groups.

**Figure 1B.**
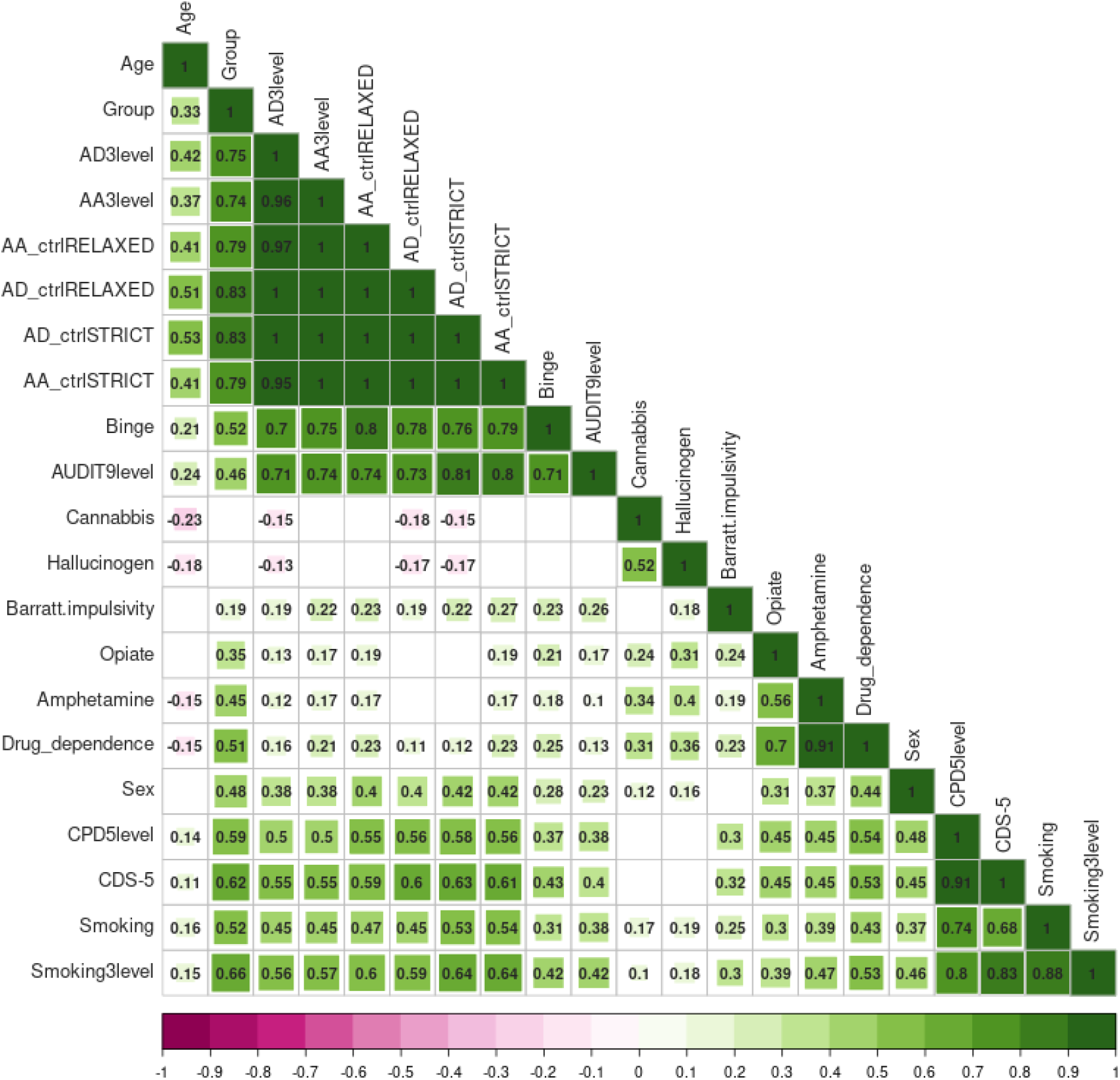
Spearman correlation plot with significant (*P*<0.05) correlations between phenotypes shown and r^2^ values reported.

The allele frequencies at *COMT* rs4680 were in line with 1000 Genomes EUR data (minor allele frequency 0.49 vs. 0.50, *P*=0.57), whereas at *HTR2A* rs6313 (0.36 vs. 0.44, *P*=0.0017), *ADH1B-ADH1C* rs1789891 (0.12 vs. 0.15, *P*=0.03), and *OPRM1* rs1799971 (0.14 vs. 0.16, *P*=0.007) we observed statistically significant differences from the 1000 Genomes EUR data (**Supplementary Table 2A**).

Across logistic and ordered logistic regression analyses, the rs4680-A at *COMT* was consistently associated with smoking status at nominal significance (OR[95% CI]=1.56[1.01-2.41], *P*=0.047) and quantity (CPD5level) (1.37[1.04-1.80], *P*=0.026) (**Table 2**). The *ADH1B* rs1789891-A showed a nominally significant association with alcohol dependence at 3-level association analysis (1.69 [1.03 – 2.76], *P*=0.038), while *HTR2A* rs6313-T was protective in this model (0.66 [0.47 – 0.92], *P*=0.0159) (**Table 2**). We did not observe any statistically significant associations with drug dependence, neither with all the drugs combined nor with any specific drug type (**Table 2**) except for *HTR2A* rs6313-T which was protective from hallucinogen use at nominal significance (0.58 [0.35 – 0.98], *P*=0.040) (**Table 2**).

## 4 Discussion

In this study with detailed phenotypic data on multiple SUDs in a Ukrainian sample, we have replicated at the strict Bonferroni-corrected threshold of significance the association of rs6313 at *HTR2A* with alcohol dependence. We show further evidence for associations at nominal significance between *ADH1B-ADH1C-*rs1789891 and alcohol dependence, *COMT-*rs4680 and smoking, and *HTR2A-*rs6313 and hallucinogen use, simultaneously controlling for other substance phenotypes in the association analyses.

The *COMT* gene (22q11.21) encodes catechol-O-methyltransferase, an enzyme that plays an important role in the metabolism of catecholamines such as dopamine, adrenaline, and norepinephrine, and is expressed in many tissues, with some predominance of RNA expression in the CNS, liver, and adrenal gland (The Human Protein Atlas, 2025; GTEx Consortium, 2025). COMT-dependent dopamine degradation is of particular importance in brain regions with low expression of the presynaptic dopamine transporter, such as the prefrontal cortex (Brodal, 2016). The rs4680 is a missense Val158Met polymorphism that is an eQTL for *COMT* itself and several other genes (GTEx Consortium, 2025); in addition to that, Met/Met (AA) genotype has been reported to lead to a 3-to-4-fold decrease in COMT enzyme activity in comparison to the wild-type GG genotype (Dawling et al. 2001).

Associations of rs4680 with smoking status, heaviness, and cessation have been previously reported in candidate gene studies (Munafò et al., 2008, 2011; Suriyaprom et al. 2013) but not in GWASs; on the other hand, GWASs have reported associations of rs4680 with various metabolites (Hysi et al. 2022; Schlosser et al. 2023; Wang et al. 2024) and internet addiction (Haghighatfard et al. 2023). Here we show evidence for the association of rs4680-A allele with smoking status and with the categorized cigarettes per day measure (CPD5level), representing smoking heaviness. This highlights a possible role of prolonged dopamine signaling in prefrontal cortex in affecting smoking behaviour. Nicotine has for a long time been shown to upregulate the release of dopamine in this area, along with the ventral tegmental area of the midbrain, nucleus accumbens, the main pathways in drug-induced reward (Benowitz, 2009). Moreover, as shown by neuroimaging, nicotine has also repaired reduced dopamine levels and working memory in preclinical studies in monkeys (Tsukada et al., 2005), and fMRI in people has shown different responses in activation of the involved brain regions to nicotine and placebo in different genotype carriers (Lee, et al. 2013).

The *ADH1B-ADH1C* locus (4q23) has in turn been consistently associated with alcohol-related variables (Frank et al. 2012; Treutlein et al. 2017; Way et al. 2015; Bach et al. 2019), as it is also in our study. The rs1789891 is an intergenic SNP in the *ADH1B-ADH1C* gene region and is an eQTL for several loci from and outside the *ADH1B-ADH1C* locus (University of Cambridge, 2025; 59). The C allele has been shown to downregulate expression of *ADH1A, ADH1C* in blood, aorta, adipose tissue, esophagus mucosa (Hibberd et al. 2020). It has been firstly associated with AD in GWASs in German samples (Frank et al. 2012; Treutlein et al. 2017). The associations of rs1789891 with AD were then replicated in candidate gene studies (Way et al. 2015). It has also been associated with relapse-free (alcohol) survival rate, ethanol intake, and craving for alcohol (Bach et al. 2019). We have observed an association of rs1789891-A with AD when analysing the 3-level AD phenotype which allowed to include all genotype-phenotype variation via using an intermediate phenotype group of people who cannot be attributed neither to people with AD nor to controls with low alcohol consumption. We might speculate that this approach increases the power to detect modest associations. Notably, we have not replicated the rs1789891-A effect on alcohol consumption reported in the UK Biobank (Alcohol intake frequency) (Canela-Xandri et al. 2018) and in a small dataset of Germans (Bach et al. 2019). Differences in the LD structure of the *ADH1B-ADH1C* region in the Ukrainian population (Edenberg 2007) and hence, the presence of another causative variant in the *ADH* region for the alcohol consumption, or insufficient statistical power may contribute to the failure to replicate this association.

The third locus, *HTR2A* (13q14.2), is a general SUD-associated locus, with reported associations spanning from smoking and nicotine addiction (Pérez-Rubio et al. 2017) to alcohol abuse (Cao et al. 2014), relapse after alcohol treatment (Jakubczyk et al. 2013) and to SUDs combined, including alcohol and heroin use (Cao et al. 2014). *HTR2A* encodes the 5-hydroxytryptamine (serotonin or 5-HT) 2A receptor which has been shown to play a role in reward system and substance dependence (Cao et al. 2014; López-Giménez et al. 2001; Kreek et al. 2005). Here, we report a protective association of coding synonymous SNP rs6313-T with 3-level AD phenotype in Ukrainians. This association is in line with previous results from other populations, including a large meta-analysis of alcohol dependence and alcohol abuse, as well as a meta-analysis of alcohol and heroin abuse, both in European and Asian samples (Cao et al. 2014). Interestingly, the CC genotype was significantly associated with increased relapse rate in alcohol treatment centers in the neighboring Poland (Jakubczyk et al. 2013).

While previously associated with heroin use, we report an association between *HTR2A*-rs6313 and hallucinogen use. Hallucinogens, such as LSD, mescaline, and psilocybin, bind with high affinity and activate the serotonin 5-HT2A receptor (López-Giménez, González-Maeso 2018). Recently, an in vitro study has shown statistically significant, although modest, effects of several non-synonymous SNPs in *HTR2A* on the efficacy and potency of four therapeutically relevant psychedelics (Schmitz et al. 2022). Accounting for this, our reported association of rs6313 with hallucinogen use seems biologically plausible, however, further studies with a significantly higher number of participants and from diverse populations are necessary to replicate this association.

We have compared the genotype frequencies in our dataset with the 1000 Genomes EUR data (2025) (**Supplementary Table A2**), and observed statistically significant discrepancies in the minor allele frequencies at *HTR2A, ADH1B-ADH1C* rs1789891, and *OPRM1* rs1799971. These might be due to the heterogeneity of the EUR superpopulation or due to the relatively small sample size of the Ukrainian dataset. However, a presence of true differences in the allelic/genotyping distributions cannot be ruled out and needs to be investigated further in a larger dataset and with inclusion of more inclusive and large reference databases.

Our study poses notable strengths. First, we collected and analysed multiple SUD phenotypes within one study. Comorbidity among various SUDs is prevalent. Epidemiological and clinical evidence suggest that polysubstance use is a standard pattern and is important to consider (Liu et al. 2018); thus, investigating substances in isolation may fail to capture the clinical reality of addiction (Cicero et al. 2020). We included patients with comorbid SUDs who are frequently excluded from such analyses (Nelson et al. 2014; Fiatal et al. 2016; Xu et al. 2024); nonetheless, to distinguish effects specific to each SUD, other substance use phenotypes were controlled for in all association analyses. Furthermore, we have considered different aspects of different substance use disorders, e.g. not only AD, but also regular consumption based on AUDIT-C scale, binge drinking and heavy drinking phenotypes, as well as very moderate and intermediate drinking patterns. We believe this approach better reflects the complexity of substance use phenotypic variation. Second, the same questionnaire and scales were applied to the cases and controls. This allowed to (i) define cases for those phenotypes that could have been defined via questionnaire responses, like smoking status or cannabis/hallucinogen use; and (ii) to select intermediate phenotypes and controls, given that some people that had no ICD-10 diagnosis and had not consulted with narcologists still had some intermediate phenotypes, mainly in regards to alcohol use. Finally, we performed the analyses on a previously understudied Ukrainian population, which have not been included in GWASs or candidate gene studies before.

This study has certain limitations as well, notably the relatively small sample size, as it includes 595 participants so far. Further participant recruitment would be highly beneficial, particularly the inclusion of more non-smoking people and more opiate drug users. We also had a limited ability to collect the data on possible social and family confounders. In addition, due to the unavailability of genome-wide data we could not assess population structure in our sample; however the people have been predominantly recruited from Kyiv region, likely limiting the heterogeneity of the sampled population. The study currently includes well-phenotyped data, genotyped at three previously reported loci, and expanding to the full genome-wide analysis would provide a more comprehensive understanding of SUD-associated loci in Ukrainians. Despite the limitations, our study has provided further evidence for three published SUD-associated loci in a previously unstudied population, with plausible biological evidence, contributing to the puzzle of SUD susceptibility in a wider population spectrum.

## Data Availability

The individual-level data that support the findings of this study are not openly available due to reasons of sensitivity and potential harm to the participants in case of data leakages or accidental identification. Some of the data may be available from the corresponding author upon a reasonable request. Data are located in a controlled access data storage.

## Acknowledgements

We are grateful to the personnel of O.M. Marzeiev Institute for Public Health NAMS Ukraine, MedLux rehab centre, Kyiv Regional Psychiatric, Narcological Medical Association in Glevakha, D.F.Chebotarev Institute of Gerontology, and T. Shevchenko Kyiv National University for support of the project at its different stages, via scientific communication on the matter, organisation of the patient recruitment, or advise on Ethical Committees documentation.

## Author contributions

Conceptualization: VB, MK, IP, OT. Methodology, Software: VB, MK. Validation: VB, ND, DO, OM. Formal analysis: VB, MK. Investigation: VB, OZ, YuB, YZ, VV, IA, ND. Resources: DO, AK, OM, OB, GZ, OT. Data Curation: VB. Writing: VB, MK, IP, OZ, YuB, YZ, VV.

Visualization: VB, MK. Supervision: MK, VB, IP. Project administration: VB, MK. OT. Funding acquisition: VB, MK, IP.

## Declaration of generative AI and AI-assisted technologies use

For function automatisation and debugging in statistical analysis, Claude Sonnet 4.0 (Anthropic) and ChatGPT-4 Turbo (OpenAI) have been used to supplement manual coding.

