## Supplementary data for "Variation at *COMT, ADH1B-ADH1C* and *HTR2A* is associated with substance use disorders in Ukrainians"

Table A1. Characteristics of the genetic variants included in the study.

| Gene | SNP | Location | Type | Major allele | Minor allele | Associated SUD-related traits according to literature - candidate gene studies | Full Reference | Associated SUD-related traits according to GWASs | Refs | Primers/probes used in the study |
|---|---|---|---|---|---|---|---|---|---|---|
| *COMT* | rs4680 | 22q11.21 | Exonic (missense) | G | A | Smoking cessation | Munafò, M. R., Johnstone, E. C., Guo, B., Murphy, M. F. G., & Aveyard, P. (2008). Association of COMT Val108/158Met genotype with smoking cessation. Pharmacogenetics and Genomics, 18(2), 121–128. https://doi.org/10.1097/FPC.0b013e3282f44daa | None; some metabolite levels | Cerezo, M., et al. (2025). The NHGRI-EBI GWAS Catalog: Standards for reusability, sustainability and diversity. Nucleic Acids Research, 53(D1), D998–D1005. https://doi.org/10.1093/nar/gkae1070 | 5'GATGGTGGATTTCGCTGGCG3' |
| | | | | | | heaviness of smoking | Munafò, M. R., Freathy, R. M., Ring, S. M., & Smith, G. D. (2011). Association of COMT Val(108/158)Met genotype and cigarette smoking in pregnant women. Nicotine & Tobacco Research, 13(2), 55–63. https://doi.org/10.1093/ntr/ntq209 | | | 5'GATGGTGGATTTCGCTGGCA3' |
| | | | | | | smoking status | Suriyaprom, K., Tungtrongchitr, R., & Harnroongroj, T. (2013). Impact of COMT Val 108/158 Met and DRD2 Taq1B gene polymorphisms on vulnerability to cigarette smoking of Thai males. Journal of Molecular Neuroscience, 49(3), 544–549. https://doi.org/10.1007/s12031-012-9844-z | | | 5'GGCCTGGTGATAGTGGGTTT3' |
| | | | | | | early onset of alcohol dependence | Nedic Erjavec, G., Nenadic Sviglin, K., Nikolac Perkovic, M., Muck-Seler, D., Jovanovic, T., & Pivac, N. (2014). Association of gene polymorphisms encoding dopaminergic system components and platelet MAO-B activity with alcohol dependence and alcohol dependence-related phenotypes. Progress in Neuro-Psychopharmacology and Biological Psychiatry, 54, 321–327. https://doi.org/10.1016/j.pnpbp.2014.07.002 ScienceDirect | | | |
| | | | | | | brain response to amphetamine | Mattay, V. S., Goldberg, T. E., Fera, F., et al. (2003). Catechol O-methyltransferase val158-met genotype and individual variation in the brain response to amphetamine. Proceedings of the National Academy of Sciences of the United States of America, 100(10), 6186–6191. https://doi.org/10.1073/pnas.0931309100 | | | |
| *ADH1B-ADH1C* | rs1789891 | 4q23 | Intergenic | C | A | Alcohol dependence | Way, M., McQuillin, A., Saini, J., Ruparelia, K., Lydall, G. J., et al. (2015). Genetic variants in or near ADH1B and ADH1C affect susceptibility to alcohol dependence in a British and Irish population. Addiction Biology, 20(3), 594–604 | Alcohol dependence | Treutlein, J., Frank, J., Streit, F., Reinbold, C. S., Juraeva, D.,et al (2017). Genetic Contribution to Alcohol Dependence: Investigation of a Heterogeneous German Sample of Individuals with Alcohol Dependence, Chronic Alcoholic Pancreatitis, and Alcohol-Related Cirrhosis. Genes, 8(7), 183. https://doi.org/10.3390/genes8070183 | 5'-ATGAGGATGCTCTCGATGTCA-3' |
| | | | | | | Relapse-free (alcohol) survival rate | Bach, P., Zois, E., Vollstädt-Klein, S., Kirsch, M., et al (2019). Association of the alcohol dehydrogenase gene polymorphism rs1789891 with gray matter brain volume, alcohol consumption, alcohol craving and relapse risk. *Addiction biology, 24*(1), 110–120. https://doi.org/10.1111/adb.12571 | | Frank, J., Cichon, S., Treutlein, J., Ridinger, M., Mattheisen, M., et al (2012). Genome-wide significant association between alcohol dependence and a variant in the ADH gene cluster. Addiction biology, 17(1), 171–180. https://doi.org/10.1111/j.1369-1600.2011.00395.x | 5'-GAAAGTCTTAAATAGAAGCAGGA-3' |
| | | | | | | Ethanol intake | Bach, P., Zois, E., Vollstädt-Klein, S., Kirsch, M., et al (2019). Association of the alcohol dehydrogenase gene polymorphism rs1789891 with gray matter brain volume, alcohol consumption, alcohol craving and relapse risk. *Addiction biology, 24*(1), 110–120. https://doi.org/10.1111/adb.12571 | | | 5'-GAAAGTCTTAAATAGAAGCAGGC-3' |
| | | | | | | Craving for alcohol | Zois, E., Vollstädt-Klein, S., Hoffmann, S., Reinhard, I., et al. (2017). Association of the alcohol dehydrogenase gene polymorphism rs1789891 with gray matter brain volume, alcohol consumption, alcohol craving, and relapse risk: ADH gene effects in alcoholism. Addiction Biology, 24(1), 110–120 | | | |
| *HTR2A* | rs6313 | 13q14.2 | Synonymous | C | T | smoking | Pérez-Rubio, G., Ramírez-Venegas, A., Díaz, V. N., García-Gómez, L., Fabián, K. E., García-Carmona, S., López-Flores, L. A., Ambrocio-Ortiz, E., Contreras-Romero, R., Alcantar-Ayala, N., Sansores, R. H., & Falfán-Valencia, R. | None | | 5`-GAGCTCAACTACGAACTCCCTA-3` |
| | | | | | | heaviness of nicotine addiction | Pérez-Rubio, G., Ramírez-Venegas, A., Díaz, V. N., García-Gómez, L., Fabián, K. E., García-Carmona, S., López-Flores, L. A., Ambrocio-Ortiz, E., Contreras-Romero, R., Alcantar-Ayala, N., Sansores, R. H., & Falfán-Valencia, R. (2017). Polymorphisms in HTR2A and DRD4 predispose to smoking and smoking quantity. *PLOS ONE, 12*(1), e0170019. | | | 5`-CCCTTCACAGGAAAGGTTGGT-3` |
| | | | | | | substance use disorders combined (alcohol + heroin) (meta-analysis) | Cao, J., Liu, X., Han, S., Zhang, C. K., Liu, Z., & Li, D. (2014). Association of the HTR2A gene with alcohol and heroin abuse. Human Genetics, 133(3), 357–365. | | | 5`Fam-TACAGTAATGACTTTAACTCCGGAGAA-3`BHQ1 |
| | | | | | | alcohol abuse (meta-analysis) | Cao, J., Liu, X., Han, S., Zhang, C. K., Liu, Z., & Li, D. (2014). Association of the HTR2A gene with alcohol and heroin abuse. Human Genetics, 133(3), 357–365. | | | 5`Hex-TACAGTAATGACTTTAACTCTGGAGAA-3`BHQ2 |
| | | | | | | Relapse after alcohol treatment | Jakubczyk, A., Klimkiewicz, A., Kopera, M., Krasowska, A., Wrzosek, M., Matsumoto, H., Burmeister, M., Brower, K. J., & Wojnar, M. (2013). The CC genotype in the T102C HTR2A polymorphism predicts relapse in individuals after alcohol treatment. *Journal of psychiatric research, 47*(4), 527–533. https://doi.org/10.1016/j.jpsychires.2012.12.004 | | | |
| *OPRM1* | rs1799971 | 6q25.2 | Missense | A | G | general substance dependence | Schwantes-An, T. H., Zhang, J., Chen, L. S., et al. (2016). Association of the OPRM1 Variant rs1799971 (A118G) with Non-Specific Liability to Substance Dependence in a Collaborative de novo Meta-Analysis of European-Ancestry Cohorts. *Behavior genetics, 46*(2), 151–169. https://doi.org/10.1007/s10519-015-9737-3 | Opioid use disorder | Xu, H., Toikumo, S., Crist, R. C., Glogowska, K., et al. | 5'CAGATGCTCAGCTCGGTCC3' |
| | | | | | | less heavy drinking days | Weerts, E. M., Wand, G. S., Maher, B., Xu, X., Stephens, M. A., Yang, X., & McCaul, M. E. (2017). Independent and Interactive Effects of OPRM1 and DAT1 Polymorphisms on Alcohol Consumption and Subjective Responses in Social Drinkers. *Alcoholism, clinical and experimental research, 41*(6), 1093–1104. https://doi.org/10.1111/acer.13384 | | Zhou, H., Rentsch, C. T., Cheng, Z., Kember, R. L., et al (2020). Association of OPRM1 Functional Coding Variant With Opioid Use Disorder: A Genome-Wide Association Study. JAMA psychiatry, 77(10), 1072–1080. https://doi.org/10.1001/jamapsychiatry.2020.1206 | 5'CCACGCACACGATGGAGTAG3' |
| | | | | | | long-term abstinence from heroin without agonist treatment | Levran, O., Peles, E., Randesi, M., da Rosa, J. C., Adelson, M., & Kreek, M. J. (2017). The μ-opioid receptor nonsynonymous variant 118A>G is associated with prolonged abstinence from heroin without agonist treatment. *Pharmacogenomics, 18*(15), 1387–1391. https://doi.org/10.2217/pgs-2017-0092 | | Deak, J. D., Zhou, H., Galimberti, M., Levey, D. F., et al. (2022). Genome-wide association study in individuals of European and African ancestry and multi-trait analysis of opioid use disorder identifies 19 independent genome-wide significant risk loci. Molecular psychiatry, 27(10), 3970–3979. https://doi.org/10.1038/s41380-022-01709-1 | 5'CTTGTCCCACTTAGATGGCA3' |
| | | | | | | drug addiction | Al-Eitan, L. N., Rababa'h, D. M., & Alghamdi, M. A. (2021). Genetic susceptibility of opioid receptor genes polymorphism to drug addiction: A candidate-gene association study. BMC psychiatry, 21, 1-14. | | Hatoum, A. S., Colbert, S. M. C., Johnson, E. C., et al. (2023). Multivariate genome-wide association meta-analysis of over 1 million subjects identifies loci underlying multiple substance use disorders. Nature. Mental health, 1(3), 210–223. https://doi.org/10.1038/s44220-023-00034-y | 5'CTTGTCCCACTTAGATGGCG3' |
| | | | | | | | | | Kember, R. L., Vickers-Smith, R., Xu, H., Toikumo, S., et al. (2022). Cross-ancestry meta-analysis of opioid use disorder uncovers novel loci with predominant effects in brain regions associated with addiction. Nature neuroscience, 25(10), 1279–1287. https://doi.org/10.1038/s41593-022-01160-z | |

**Table A2. Comparison of the genotype frequencies in the Ukrainian controls outside rehab centres with 1000 Genomes EUR data.**

| | | | | | | | | | | |
|---|---|---|---|---|---|---|---|---|---|---|
| *HTR2A* rs6313 (C/T) | | | | | | | | | | |
| Ctrls | | | | | https://useast.ensembl.org/Homo_sapiens/Variation/Population?db=core;r=13:46895305-46896305;v=rs6313;vdb=variation;vf=813010869 | | | | | |
| Genotypes | N observed | Alleles | N observed | freq alleles observed | Genotypes 1000 Genomes EUR | N 1000 Genomes EUR | 1000 G N normalised for sample size (=exp) genotypes | Alleles | N 1000 Genomes EUR | freq alleles 1000 Genomes EUR |
| CC | 103 | C | 336 | 0.636363636 | CC | 166 | 87.1252485089463 | C | 567 | 0.564 |
| CT | 130 | T | 192 | 0.363636364 | CT | 235 | 123.339960238569 | T | 439 | 0.436 |
| TT | 31 | | | | TT | 102 | 53.5347912524851 | | | |
| | sample (dataset) | 264 | | | sample 1000G | 503 | | | | |
| | (o-e)2/e | 2.892476512 | | | | | | | | |
| | | 0.359624971 | | | | | | | | |
| | | 9.485734509 | | | | | | | | |
| | X2 | **12.73783599** | | | | | | | | |
| | p | **0.001714** | | | | | | | | |
| *ADH1B-ADH1C* rs1789891 | | | | | | | | | | |
| Ctrls | | | | | https://useast.ensembl.org/Homo_sapiens/Variation/Population?db=core;r=4:99328762-99329762;v=rs1789891;vdb=variation;vf=261294930 | | | | | |
| Genotypes | N observed | Alleles | N observed | freq alleles observed | Genotypes 1000 Genomes EUR | N 1000 Genomes EUR | 1000 G N normalised for sample size (=exp) genotypes | Alleles | N 1000 Genomes EUR | freq alleles 1000 Genomes EUR |
| AA | 0 | A | 64 | 0.121673004 | AA | 13 | 6.79721669980119 | A | 148 | 0.147 |
| AC | 64 | C | 462 | 0.878326996 | AC | 122 | 63.7892644135189 | C | 858 | 0.853 |
| CC | 199 | | | | CC | 368 | 192.41351888668 | | | |
| | sample (dataset) | 263 | | | sample 1000G | 503 | | | | |
| | (o-e)2/e | 6.7972167 | | | | | | | | |
| | | 0.000696191 | | | | | | | | |
| | | 0.225460943 | | | | | | | | |
| | X2 | **7.023373834** | | | | | | | | |
| | p | **0.02985** | | | | | | | | |
| *COMT* rs4680 (Val158Met) G/A | | | | | | | | | | |
| Ctrls | | | | | https://useast.ensembl.org/Homo_sapiens/Variation/Population?db=core;r=22:19963248-19964248;v=rs4680;vdb=variation;vf=1093709672 | | | | | |
| Genotypes | N observed | Alleles | N observed | freq alleles observed | Genotypes 1000 Genomes EUR | N 1000 Genomes EUR | 1000 G N normalised for sample size (=exp) genotypes | Alleles | N 1000 Genomes EUR | freq alleles 1000 Genomes EUR |
| AA | 60 | A | 246 | 0.488095238 | AA | 133 | 66.6322067594433 | A | 503 | 0.5 |
| AG | 126 | G | 258 | 0.511904762 | AG | 237 | 118.735586481113 | G | 503 | 0.5 |
| GG | 66 | | | | GG | 133 | 66.6322067594433 | | | |
| | sample (dataset) | 252 | | | sample 1000G | 503 | | | | |
| | (o-e)2/e | 0.66013372 | | | | | | | | |
| | | 0.444447241 | | | | | | | | |
| | | 0.005998381 | | | | | | | | |
| | X2 | 1.110579342 | | | | | | | | |
| | p | 0.5739 | | | | | | | | |
| *OPRM1* rs1799971 | | | | | | | | | | |
| Ctrls | | | | | https://useast.ensembl.org/Homo_sapiens/Variation/Population?db=core;r=6:154039162-154040162;v=rs1799971;vdb=variation;vf=406243391 | | | | | |
| Genotypes | N observed | Alleles | N observed | freq alleles observed | Genotypes 1000 Genomes EUR | N 1000 Genomes EUR | 1000 G N normalised for sample size (=exp) genotypes | Alleles | N 1000 Genomes EUR | freq alleles 1000 Genomes EUR |
| AA | 182 | A | 411 | 0.85625 | AA | 353 | 168.429423459245 | A | 843 | 0.838 |
| AG | 47 | G | 69 | 0.14375 | AG | 137 | 65.3677932405567 | G | 163 | 0.162 |
| GG | 11 | | | | GG | 13 | 6.20278330019881 | | | |
| | sample (dataset) | 240 | | | sample 1000G | 503 | | | | |
| | (o-e)2/e | 1.093398908 | | | | | | | | |
| | | 5.161193484 | | | | | | | | |
| | | 3.710155095 | | | | | | | | |
| | X2 | **9.964747487** | | | | | | | | |
| | p | **0.006858** | | | | | | | | |

Figure A1. Distributions of the scale variables.

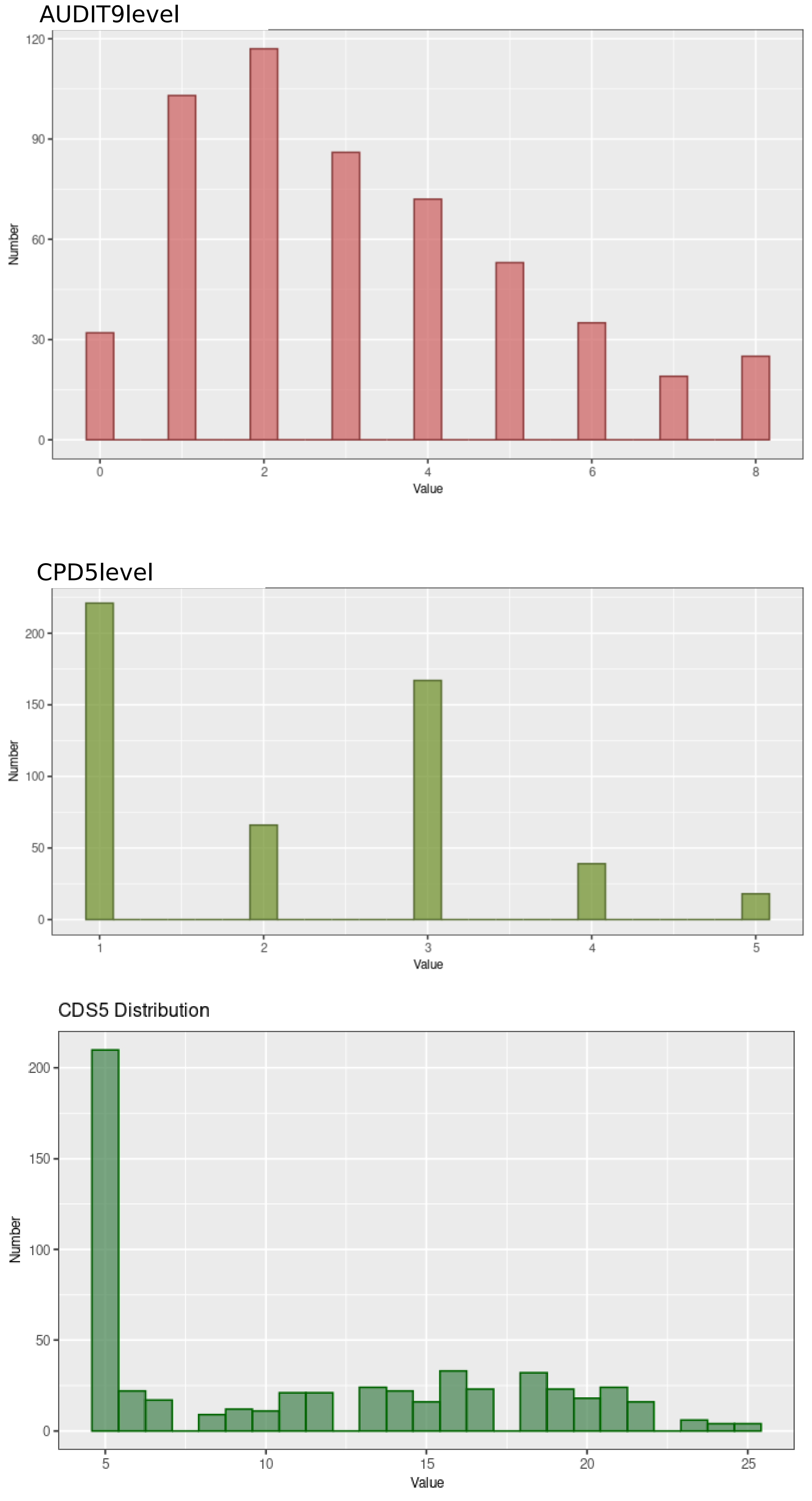
